# Genetic Association of Glutathione S-Transferase Omega 2 *(GSTO2)* Gene Variant (rs156697) with Chronic Kidney Disease Patients of Pakistani Origin

**DOI:** 10.1101/2024.10.01.24313369

**Authors:** Abeer Afzal, Rashid Saif, Asia Atta, Saghir Ahmad Jafri, Syeda Komal Abbas Naqvi

## Abstract

Chronic Kidney Disease (CKD) poses a significant global health challenge. The kidneys can be compromised by internal diseases, environmental influences and various genetic factors. Among the genetic factors, particularly, *GSTO2* gene plays a crucial role in cellular processes and enzyme performance, which can lead to interstitial kidney damage and contribute to CKD. The current study investigates the genetic association between the *GSTO2* gene variation (rs156697) and CKD in the Punjabi community. This study examined the association between the *GSTO2* gene locus (c.424A>T) and CKD in a Punjabi population using ARMS-PCR genotyping. A total of 50 samples were analyzed, revealing 19 homozygous wild-type, 25 heterozygous and 6 homozygous mutants. PLINK data analysis toolset indicated an insignificant genotypic association with CKD phenotype with observed *p-value* of 0.7177 and an odds ratio of 0.85. Similarly, the alternative allele frequency of 0.34 and 0.375 was also observed in CKD and normal controls respectively. In the future, further studies with more genetic markers along with functional studies with large sample data may be performed for robust statistical evidence of the association of subject gene with CKD patients in Pakistan.

## Introduction

Chronic kidney disease (CKD) has emerged as a significant global public health challenge, characterized by rising morbidity and mortality rates. The increasing prevalence of metabolic diseases has escalated the risk of CKD across populations, yet the primary risk factors driving the progression of renal disease remain poorly understood. Genetic factors play a crucial role in CKD development, identifying specific gene-environment interactions that directly correlate with CKD risk remains challenging (Bikbov et al., 2020). Genome-wide association studies (GWAS) have advanced our understanding of genetic influences on common diseases, including CKD, by examining the relationship between gene variations and disease expression (Melzer et al., 2008).

Recent GWAS have identified associations between specific genetic variants and CKD-related phenotypes such as estimated glomerular filtration rate (eGFR), albumin levels, and serum creatinine (SCr) (Gudbjartsson et al., 2010). Notable loci linked to eGFR include SHROOM3, GATM-SPATA5L1, and UMOD, while other genes such as STC1 and CST9 have been associated with eGFR based on cystatin C (eGFRcys) in an Icelandic population. Additional genes implicated in CKD and renal function include VEGFKA, PRKAG2, DAB2, ATXN2, DACH1, ALMS1, UBE2Q2, GCKR, TFDP2, SLC34A1, SLC7A9, and PIP5K1B (Kottgen et al., 2022). Studies have also identified genes linked to albuminuria in African Americans, such as MYH9 and APOL1, highlighting significant genetic and environmental variations among populations (Freedman et al., 2009).

There is growing evidence that genetic differences contribute significantly to disease susceptibility and progression (Pattaro et al., 2016). Glutathione S-transferases (GSTs), including the GST Omega 2 (*GSTO2*) gene, are phase II detoxification enzymes involved in cellular defense against oxidative stress and regulation of inflammation. Variants in the *GSTO2* gene have been linked to various diseases, including cancer, neurodegenerative disorders, and cardiovascular disease, due to their role in oxidative stress regulation (Prysyazhnyuk et al., 2021). The rs156697 mutation in *GSTO2* has been specifically associated with CKD in diverse populations (Corredor et al., 2020). Punjab, with its unique genetic and ethnic diversity, offers a valuable context for studying genetic variations contributing to CKD. The region has a high prevalence of CKD, with significant socioeconomic implications (Imran et al., 2015). This study aims to explore the association between the *GSTO2* gene variant rs156697 and CKD in a Punjabi community.

## Material and Methods

### Sample collection and DNA extraction

A total of 50 samples, comprising 25 CKD cases and 25 controls, were collected. The CKD cases included both genders, aged 28-72 years, with serum creatinine (SCr) levels ranging from 0.7 to 15.9 mg/dL and glomerular filtration rates (GFR) between 3 and 118 mL/min/1.73 m^2^. These samples were collected from various hospitals in Lahore, Pakistan, between September and October 2024, to investigate the association of the *GSTO2* gene SNP (rs156697) at position C.424 A>T with CKD. Blood samples were drawn into EDTA-K3 vacutainers and stored at 4°C. DNA was extracted from each sample using the GDSBio (https://www.gdsbio.com/en/) genomic DNA extraction kit, following the manufacturer’s guidelines.

### Primer designing

Based on the *GSTO2* Gene SNP (rs 156697) transcript ID: NM_183239.2 in Homo sapiens at location position C.424 A>T, five PCR primers were designed using NetPrimer software. Three of these were ARMS-PCR primers used to amplify both normal and mutant alleles, labeled as reverse normal (RN) and reverse mutant (RM), with a mismatch inserted at the fourth nucleotide position from the 3’ end to enhance accuracy and efficacy. A forward common primer (FC) was also included. Additionally, two more primers were designed to produce an amplicon of 618 bp, serving as an internal control (IC). Table 1 provides the sequences and other attributes of these primers.

**Table 1.**
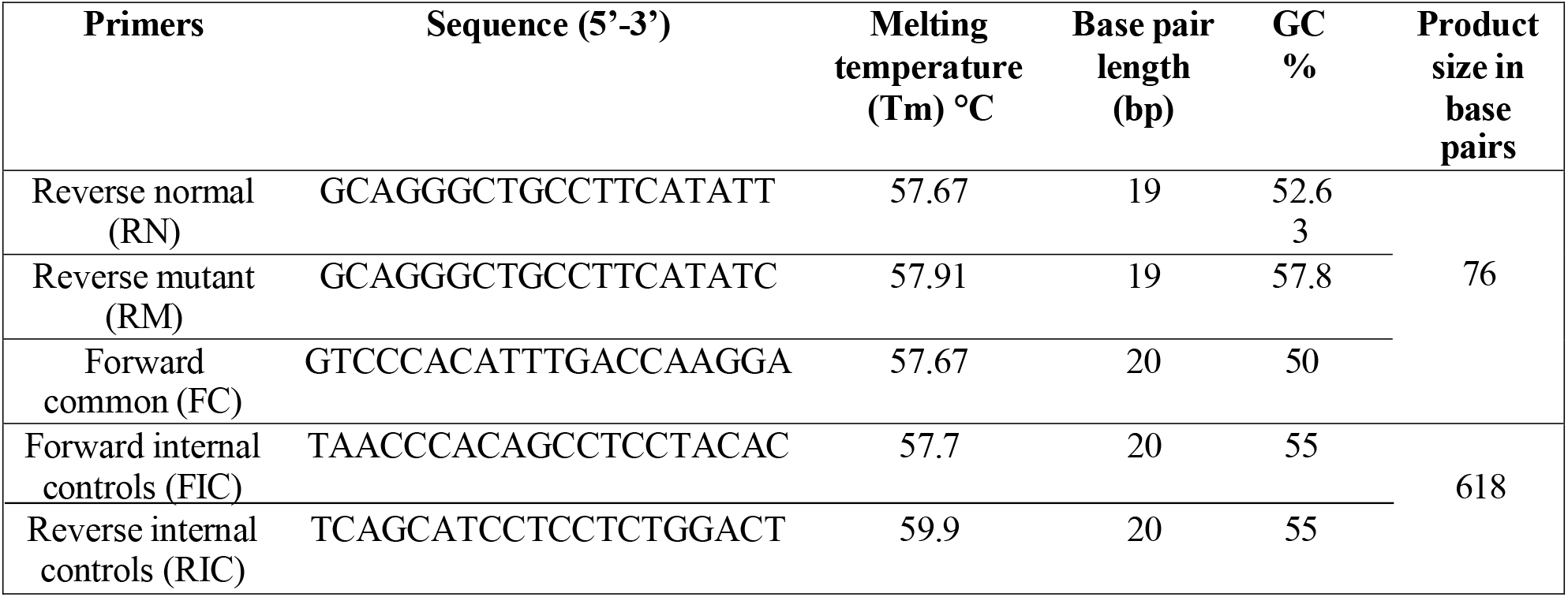
Details of primers for ARMS-PCR.

### PCR amplification

The ARMS-PCR reaction was conducted using a SimpliAmp thermal cycler from Applied Biosystems. Each sample underwent two independent reactions: one with a common forward primer and a normal ARMS reverse primer, and the other with a common forward primer and a mutant ARMS reverse primer. To ensure the fidelity of the PCR, internal control primers were included in each tube, amplifying a targeted region of known size. The reaction mixture, totaling 16 μL, consisted of the following components: 2 μL of genomic DNA (extracted at a concentration of 50 ng/μL), 10 mM of each primer (forward common, N-reverse or M-reverse primers, forward and reverse internal control primers), 0.05 IU of Taq polymerase, 2.5 mM MgCl2, 2.5 mM dNTPs, 1x Taq buffer, and molecular grade water. The PCR protocol included an initial denaturation at 95°C for 5 minutes, followed by 30 cycles of denaturation at 95°C for 30 seconds, annealing at 60°C for 30 seconds, and extension at 72°C for 45 seconds, with a final extension at 72°C for 10 minutes, and storage at 4°C.

### Statistical analysis

PLINK data toolset was used to evaluate the Hardy Weinberg Equilibrium (HWE) using equation (2 + 2 + 2 = 1) to check if sampled population is in accordance with HWE or not and Chi-Square association to find the association of subject variant with CKD after screening the rs156697A>T locus in total of fifty samples. Descriptive statistics were used to provide quantitative factors such age as Mean ± SD, whereas qualitative variables like gender, health history, diagnoses, and family health history were represented as percentages. A threshold of p ≤ 0.05 was considered statistically significant for all tests conducted within the scope of this research.

## Results

### Data Analysis

The objective of the studies was to analyse the CKD prevalence within the Punjab population. The sample size included individuals from 9 divisions out of 11 divisions of Punjab, which is about 81.81% of the all divisions. Figure 1 shows the population distribution of the sample.

**Figure 1.**
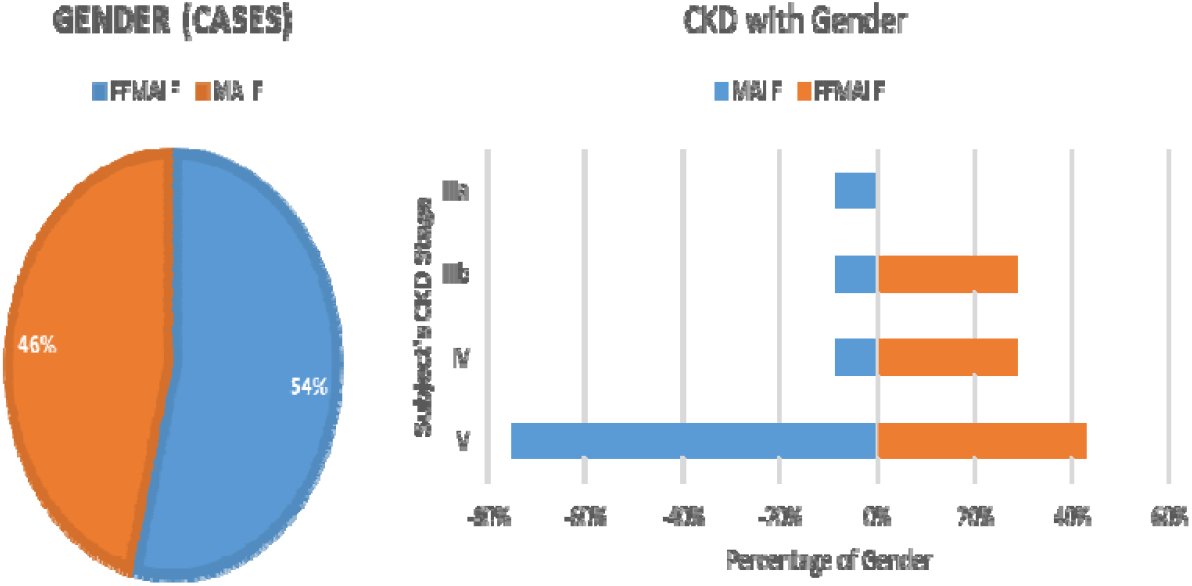
Gender distribution and CKD severity according to gender

**Figure 2.**
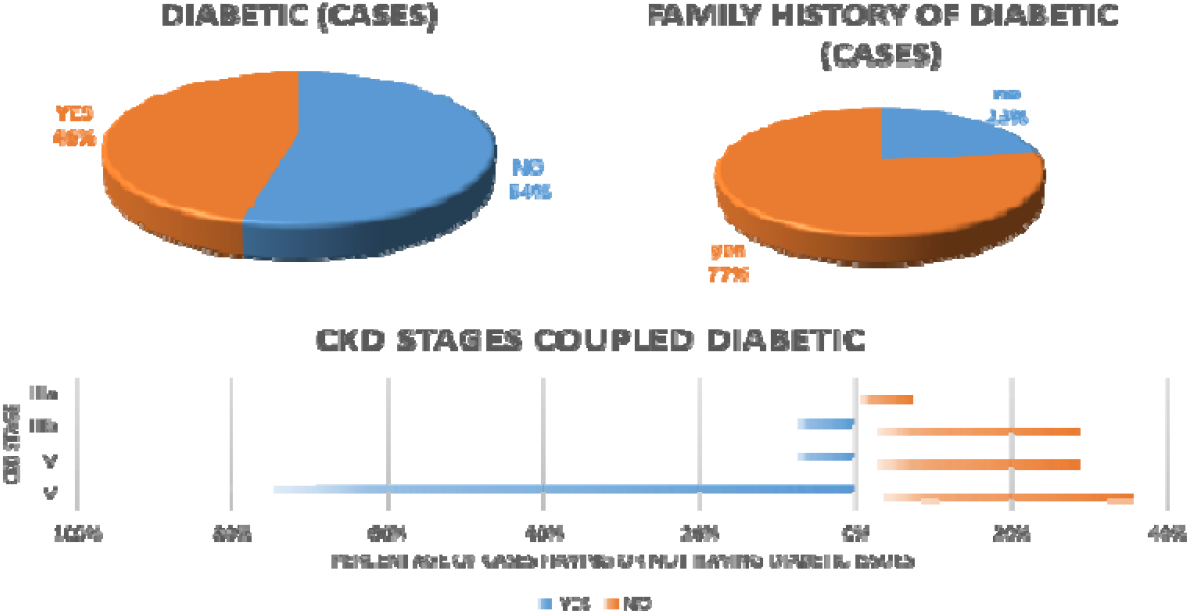
Diabetic distribution and CKD severity

**Figure 3.**
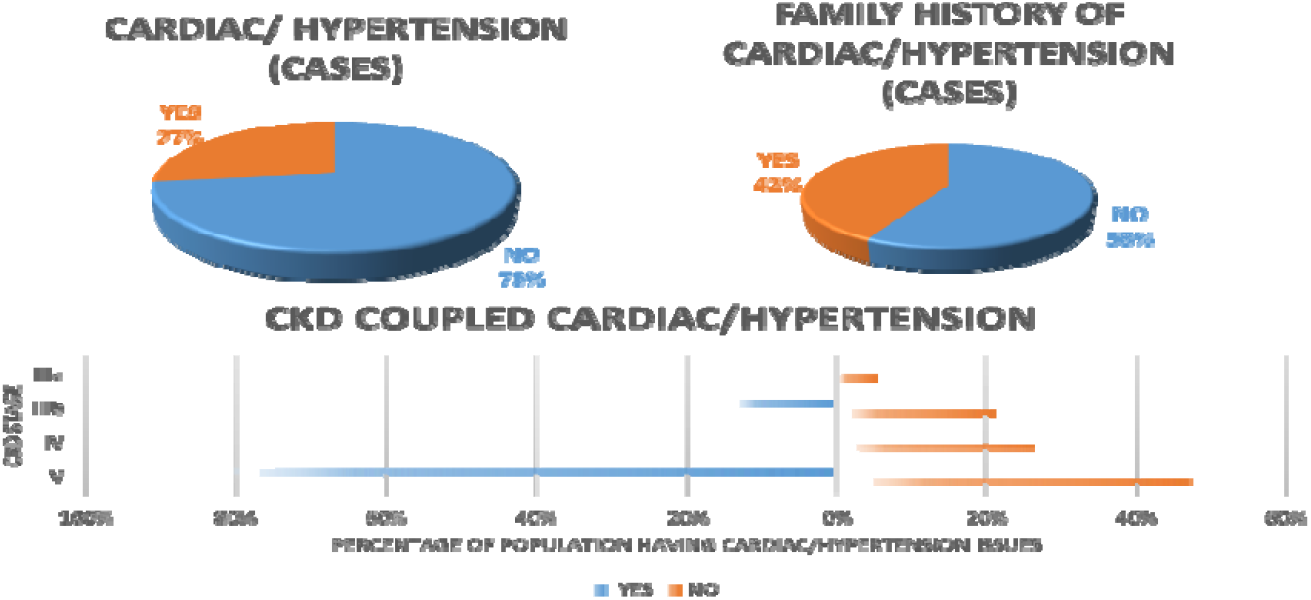
Diabetic distribution and CKD severity

**Figure 4.**
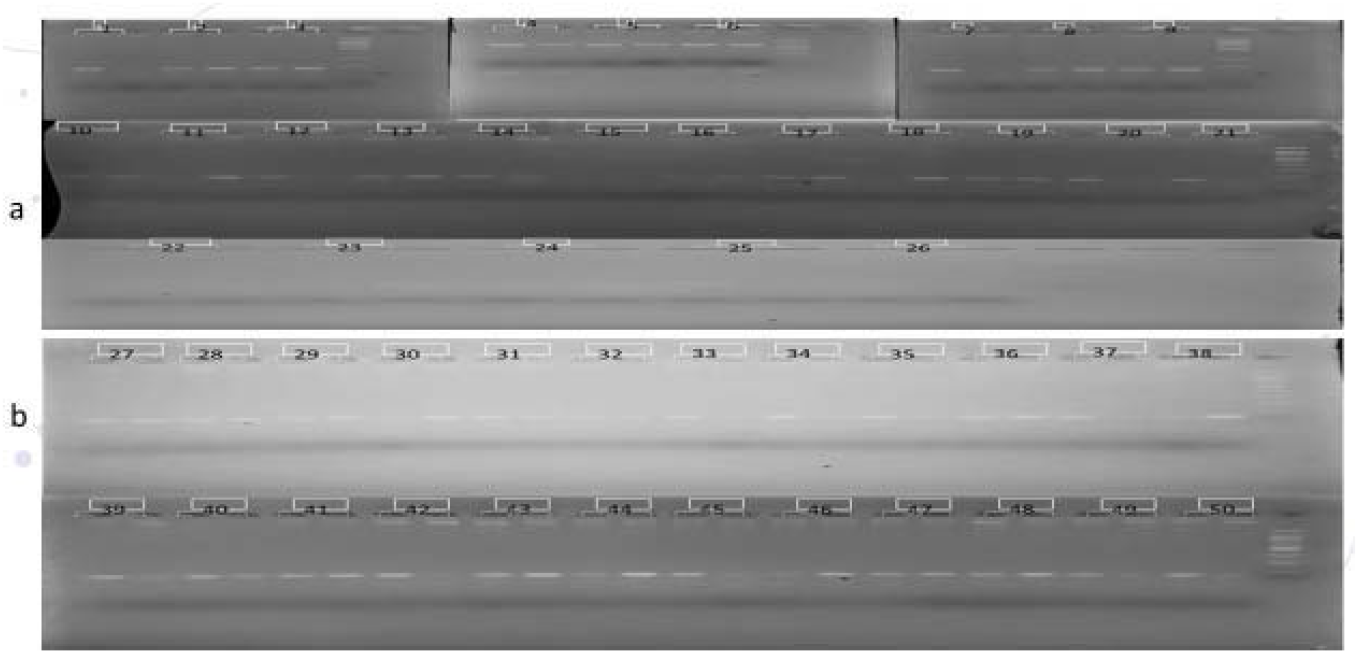
Genotyping of 26 CKD cases (a) and 24 controls (b).

The average age of total sample was about 47 years having maximum values of 72 and minimum age of 28 years, women in sample has slightly higher age than men group in total sample. The cases group has average age of 47 years, where female has 46 and men have 49 years of average age in Cases group as shown in table 2.

**Table 2:** Descriptive statistic of age according to sample size and gender.

| SAMPLE SIZE |  | Maximum | Minimum | Mean | Median | Mode | Std. deviation | Range |
| --- | --- | --- | --- | --- | --- | --- | --- | --- |
| (Control + Cases) | sample | 72 | 28 | 46.94 | 44 | 35 | 12.76 | 44 |
|  | FEMALE | 65 | 30 | 47.24 | 45 | 35 | 11.03 | 35 |
|  | MALE | 72 | 28 | 46.64 | 36 | 35 | 14.51 | 44 |
| Cases | CASES | 65 | 30 | 47.18 | 45 | 35 | 12.77 | 35 |
|  | FEMALE | 65 | 30 | 46.07 | 42.5 | 36 | 12.22 | 35 |
|  | MALE | 64 | 35 | 49.08 | 51 | 35 | 9.3 | 29 |

The female population account about 54%, while men population is 46% in cases, however higher population of men were severely affected (stage V) than females.

The non-diabetic population is higher than diabetic population in both total sample and cases, which account 33 out of 50 and 14 out of 26 respectively. About 46% of population has both diabetes and CKD, 77% of cases had family history of diabetes. it showed that more than 80% of total diabetic patient ar suffering from CKD stage V.

The 27% of cases sample reported cardiac/hypertension, while 42% have family history of cardiac/hypertension. Further more than 80% of cardiac patient have end stage of CKD.

### Variant genotyping

The current study focused on the relationship between the *GSTO2* gene variant (rs156697) and chroni kidney disease (CKD) among the Punjabi population. A genotyping technique was used on a cohort of 50 samples, with 26 cases and 24 controls. Twelve of the CKD patients had a heterozygous genotype, eleven had a homozygous wild-type genotype, and three had a homozygous mutant genotype. Similarly, 13 individuals in the control cohort had a heterozygous genotype, 8 had a homozygous wild-type genotype, and 3 had a homozygous mutant genotype. Following a thorough study of the sampled population, it wa determined that 25 individuals (50%) held a heterozygous genotype, 21 (38%) exhibited a homozygou wild-type genotype, and 6 (12%) were homozygous mutant genotype.

## Discussion

Familial clustering of CKD and kidney-related indicators implies that genetic or shared environmental variables play a role in disease pathogenesis (Wuttke et al. 2019). Females have a higher prevalence of pre-dialysis CKD than males (Tomlinson and Clase 2019), males experience a faster deterioration in kidney function (Carrero et al., 2018). For Pakistani population highest incidence of CKD among senior individuals over the age of 50 (Alam et al. 2014). CKD and diabetes are inextricably related, with diabetes being identified as a major etiological factor leading to CKD worldwide (Fenta, 2023), Diabetes is also the primary cause of end-stage renal illness in Pakistan’s metropolitan regions (Alemu et al., 2020). Bidirectional link is highlighted by the significant frequency of hypertension across CKD stages ((Burnier & Damianaki, 2023). Although 12% of the cases population had the homozygous mutant genotype linked to an increased risk of chronic kidney disease (CKD) or disease progression, this connection is statistically insignificant as a p-value (P) of 0.7177, showing a lack of significant relationship between this variation and the phenotype. SNPs rs156697 in *GSTO2* genes, showed a strong correlation with Spanish population under controlled the dominant model (Corredor et al. 2020). A notable link between the *GSTO2* rs156697 polymorphism and diabetic nephropathy when studying the diabetes association with numerous gene variations for Bosnia and Herzegovina population (Pavlovic et al., 2023).

## Conclusion

The presence of the *GSTO2* gene variant (rs156697) in the patient group is an interesting finding, but that lacks statistical significance. This might be attributed to a variety of variables, including the limits imposed by the current sample size, intrinsic differences within the examined group, or the impact of other genetic determinants. Future study recommendations include conducting larger-scale studies including varied cohorts, encouraging collaboration across research institutions, and investigating new genetic variations and environmental factors.

## Supporting information

Ethical Statement

## Data Availability

N/A

## Acknowledgement

The authors are grateful to Mr. Ahmad Saim for his support in sample collection.

## Ethical statement

The samples were obtained with consent. The study protocol was reviewed and approved by the Institutional Review Board at NUR International University, in compliance with their Ethics Committee’s regulations.

