## Supplementary figures and images for "Genetic Association of Glutathione S-Transferase Omega 2 *(GSTO2)* Gene Variant (rs156697) with Chronic Kidney Disease Patients of Pakistani Origin"

### Ethics Statement-NIU.docx

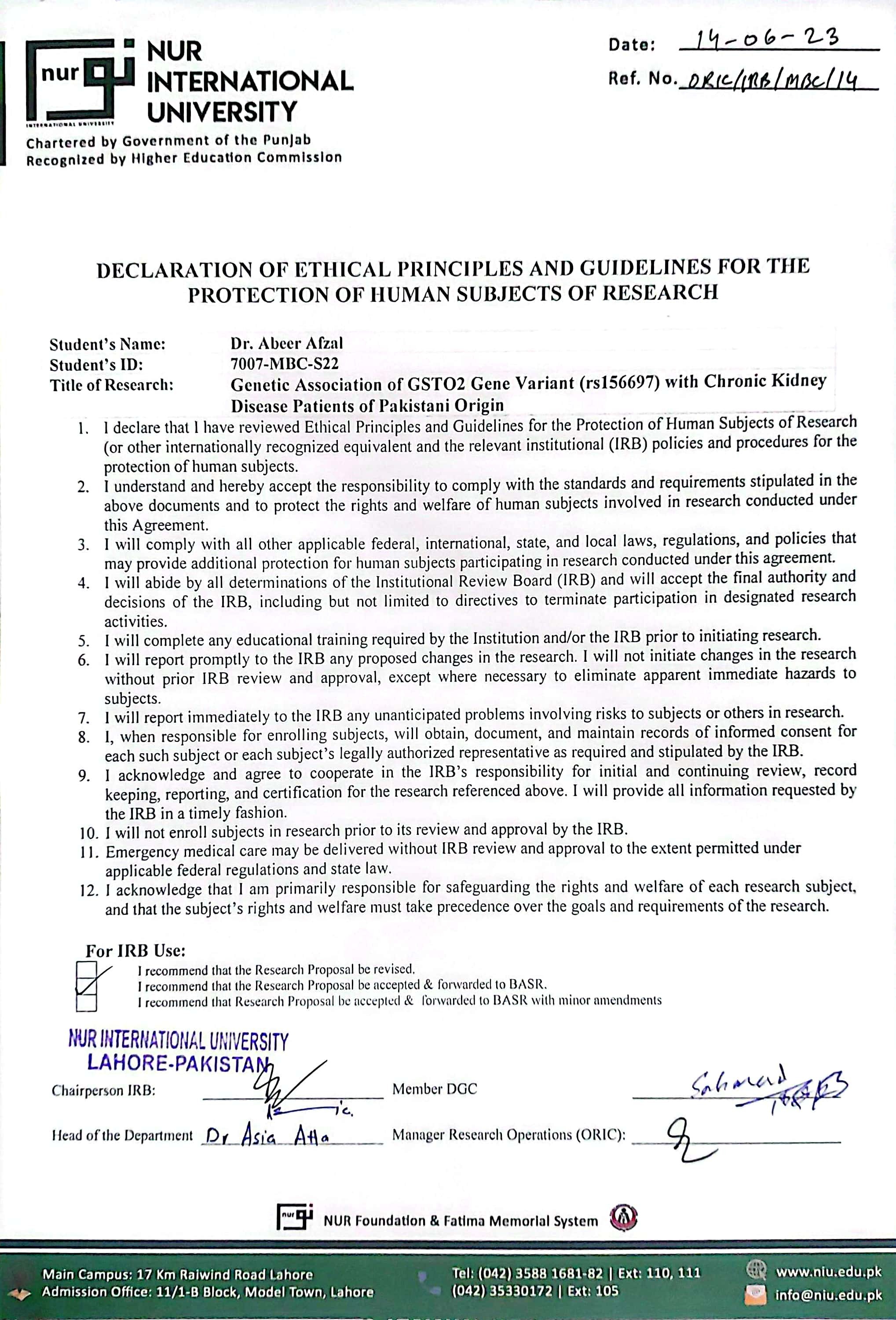
